# Disease stages and therapeutic hypotheses in two decades of neurodegenerative disease clinical trials

**DOI:** 10.1101/2022.06.16.22276513

**Authors:** Meredith A. Mortberg, Sonia M. Vallabh, Eric Vallabh Minikel

## Abstract

**Background:** Neurodegenerative disease is increasing in prevalence and remains without disease-modifying therapies, and most trials of new drugs fail. Proposed solutions include aiming upstream: targeting molecular root causes of disease and testing therapies earlier, even at pre-symptomatic stages. We sought to understand what disease stages were eligible to enroll in neurodegenerative disease clinical trials in recent years and what molecular targets were tested in these trials.

**Methods:** We combined automated analysis and manual curation of trial registrations from ClinicalTrials.gov for Alzheimer’s disease, Parkinson’s disease, frontotemporal dementia / amyotrophic lateral sclerosis, and Huntington’s disease.

**Findings:** 3,241 trials from 2000-2020 were curated. Industry-sponsored drug trials, a minority (34%) of trials but a majority (61%) of patient-years, were more likely to complete, to have specified phase, and to have placebo or standard-of-care control arms. The mean number of inclusion and exclusion criteria more than doubled over this period, and eligible score ranges shrank. Trials have shifted towards less severely impaired participants, but only 2.7% of trials were open to pre-symptomatic individuals and these were depleted for industry sponsors (OR = 0.32) and for drug trials (OR = 0.59), instead being enriched for behavioral interventions (OR = 3.1). 16 novel, genetically supported therapeutic hypotheses have been tested in drug trials, with a mean lag of 13 years from genetic association to first trial. Such trials comprised just 18% of patient-years.

**Interpretation:** Eligibility criteria for trials have shifted towards earlier, milder disease stages but are still overwhelmingly focused on symptomatic patients, particularly for industry-sponsored drug trials. Drugs targeting disease genes supported by human genetics comprise a small fraction of drug development effort, and their success may be hindered by a focus on symptomatic patients.

## Introduction

Neurodegenerative disease is on the rise globally due to aging populations^1^, highlighting a need for effective therapeutic interventions. Developing new drugs is incredibly difficult, with only 8-14% of all drug-indication pairs that enter clinical trials ultimately succeeding^2–4^. Success has been particularly limited in adult-onset neurodegenerative diseases, for which no disease-modifying drug yet exists. Patients, scientists, regulators, and public health experts have called for prioritizing preventive approaches to neurodegeneration, citing potential benefits to quality of life and alleviation of economic burden, as well as improved prospects for success if intervention against core molecular drivers of disease occurs before downstream pathology takes hold^1,5–9^. Yet there exist considerable barriers to achieving prevention, including the potential cost and duration of trials, need for deeper validation of biomarker endpoints, and the requirement for careful protections and counseling of at-risk subjects in trials^5,7,10^.

In this study, we sought to understand the disease stages and therapeutic hypotheses studied in clinical trials conducted in neurodegenerative diseases to date. Current drug development pipelines have been catalogued elsewhere^11–14^, but we found that none of these reports answered our key questions. First, experimental drugs are often broadly categorized as “disease-modifying” if the hypothesis is one of disease modification, regardless of the quality of that hypothesis. We wished to examine the share of drugs with molecular targets underpinned by human genetic associations, a factor shown empirically to double success rates in drug development^15,16^. Second, while reports sometimes categorize trials as preventive versus symptomatic, we wished to examine more quantitative metrics of disease stage or severity. Third, while snapshots of the present pipeline are illuminating, we sought a longitudinal view of drug discovery over the past two decades. Finally, prior studies did not make their full datasets publicly available to facilitate re-analysis. Therefore, combining automated annotation and deep manual curation of clinical trial registrations from ClinicalTrials.gov, we set out to map the landscape of clinical trials and their therapeutic interventions and to test correlations and trends over the past two decades.

## Methods

### Study design

Our goals were to characterize clinical trials across major neurodegenerative disease indications, identifying correlations and temporal trends, particularly with regards to disease stages and molecular targets of drugs. We chose ClinicalTrials.gov as a data source because i) it is publicly available, allowing us to release our curated dataset and source code and thus make our analyses fully reproducible, ii) it has existed since 2000, thus providing two decades of data, and iii) compliance with Food and Drug Administration Amendments Act (FDAAA) mandates have made it a fairly comprehensive listing of trials^17,18^. Trial results are not always posted in a timely fashion^19^, but our analysis focused on the design, not results, of trials. We determined that the ClinicalTrials.gov data, downloadable in XML format, necessitated a hybrid approach using both scripting and manual curation. Some variables, such as study start and end dates, enrollment, and phase are captured in specific fields and can be trivially extracted with scripts. Other variables require manual curation either because they are described using variable diction within free text fields (eligibility criteria), contain large numbers of synonyms, typos, and qualifiers (intervention names), or require cross-referencing to diverse external data sources (drug target). We therefore used a Python script to extract fields of interest into separate trials and interventions tables which were manually curated in Google Sheets.

### Search strategy and selection criteria

We included 4 major neurodegenerative diseases: Alzheimer’s disease (AD), Parkinson’s disease (PD), frontotemporal dementia / amyotrophic lateral sclerosis (FTD/ALS) and Huntington’s disease (HD). On April 16, 2020, we searched ClinicalTrials.gov for alzheimer OR huntington OR parkinson OR als OR amyotrophic OR frontotemporal OR ftd OR ftld with start date on or before March 31, 2020, yielding 4,542 NCT identifiers. Full ClinicalTrials.gov registration data for these trials were then pulled from https://clinicaltrials.gov/AllPublicXML.zip on April 16, 2020 for manual curation as described below.

### Trial curation

Because our research question centered on patient populations and therapeutic hypotheses, we sought to include all trials that tested an intervention hypothesized to modify the patient’s disease or symptoms, in a disease-relevant population (Figure 1). We excluded trials lacking a therapeutic intervention, such as studies of biomarkers, diagnostics, patient data, or imaging agents, where the goal was to evaluate diagnostic or prognostic value rather than to confer therapeutic benefit. We also excluded trials that enrolled only healthy volunteers, did not study a neurodegenerative disease, or targeted the intervention to caregivers rather than patients. Finally, we excluded trials where data were incomplete or contained errors. We performed internet searches to identify the sector to which trial sponsors belonged (industry or “other” including academia, government, non-profits). We defined disease stage 0 (“at-risk”) as individuals at risk of disease due to genotype, age, or other risk factors. Generalizing FDA Alzheimer’s guidance^7^ across diseases, we defined stage 1 (“molecular”) as individuals with molecular evidence of disease pathology, stage 2 (“detectable”) as individuals without functional impairment but where a sensitive neuropsychological test could discern disease-related phenotypic changes, stage 3 (“mild”) as indicating mild detectable functional impairment not yet meeting criteria for disease diagnosis, and stage 4 (“diagnosed”) as individuals diagnosed with dementia or other changes (such as motor impairment) meeting diagnostic criteria for neurodegenerative disease. For each trial, we manually curated the earliest and latest disease stage of patients eligible to enroll based on reading the inclusion and exclusion criteria. We also manually extracted the eligible score ranges on tests from these criteria, prioritizing the Mini-Mental State Examination (MMSE)^20^ and Hoehn and Yahr^21^ in instances where more than one test was used. For trials that specified only a minimum score, we inferred the maximum to be the maximum possible score; and vice versa. Trial duration was calculated as study completion date minus start date. Patient-years of enrollment were calculated as trial duration times enrollment, though we acknowledge this is an imperfect approximation, as in reality participants accrue (and withdraw) gradually rather than all participants being on board for a trial’s full duration. We used only enrollment numbers described as “Actual”; patient-years for “Anticipated” enrollment values were set to missing. The number of inclusion/exclusion criteria was calculated by counting the total number of items in numbered or bulleted lists under the headings “Inclusion Criteria” and “Exclusion Criteria”, or manually curated for 75 trials without numbered/bulleted lists.

**Figure 1.**
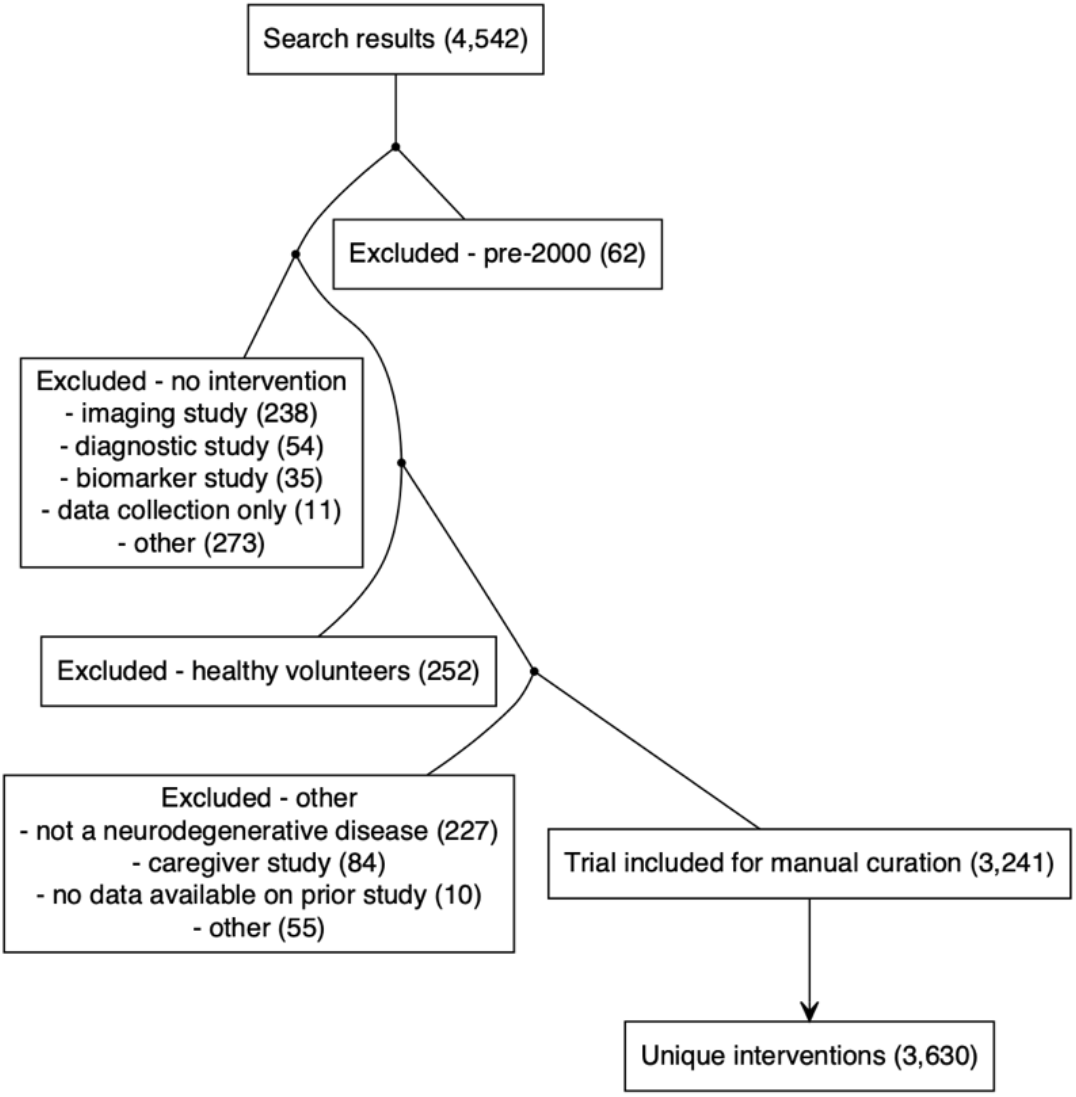
Flowchart of trial inclusion and curation. Trials returned by our search strategy were sequentially excluded based on launch year, lack of therapeutic intervention, patient population, or other. See Methods for explanation.

### Intervention curation

Intervention arms extracted included trials were converted to lower case and stripped of invalid text characters, yielding unique values for curation. The “intervention type” in ClinicalTrials.gov is specified inconsistently even for the exact same intervention (for instance, “deep brain stimulation” was categorized alternatively as “device”, “procedure”, “behavioral” or “other”), so we manually assigned every intervention to an intervention class: drug, device, procedure, behavioral, placebo, other, and none. We defined “drugs” as molecular interventions regardless of modality (synthetic drug, biological) or regulatory status (experimental, approved/repurposed, supplement). We considered as “behavioral” any interventions intended to alter patient behavior (exercise, diet, drug compliance) even if they utilized a device. We classified as “procedures” surgery, acupuncture, cell transplants, radiation, and changes in care protocols. In contrast, intervention arms that consisted solely of blood draws, lumbar punctures, or other events defined as “procedures” by Institutional Review Boards but not intended to confer therapeutic benefit to the patient, were classified as “none”. We classified as “placebo” explicit placebo arms as well as standard of care, normal saline, no treatment, and similar variations. In addition, often, a single unique intervention name value (“donepezil”) would appear across dozens of trials, where sometimes it was the active therapeutic agent being tested, while at other times it served as the standard-of-care arm. For trials with >1 intervention arm listed, if any arm was an experimental intervention while another arm was a drug that was already FDA approved for the disease in question in the year the trial occurred, we counted the latter as a placebo arm. Names of intervention arms often included information such as dose level (“riluzole 50 mg”) or formulation (“rosiglitazone xr (extended release) oral tablets”), the same drug would be assigned various generic names assigned at different stages of development (“ly3002813” is “donanemab”), and commercial names, typos, and other variations also occurred. We therefore assigned for each drug intervention arm the best generic name for the molecular entity being tested. For the 27 drugs approved by FDA for treatment of these 4 neurodegenerative diseases, years of initial approval were extracted from Drugs@FDA database searches. For the purposes of classifying experimental drugs (those not approved for these neurodegenerative diseases) as novel versus repurposed, we considered as “approved” any drug with full marketing approval in any jurisdiction worldwide. We considered only full approvals, thus aducanumab, which received Accelerated Approval in 2021, was classified as experimental. We searched several data sources to identify any available evidence as to the molecular target of each drug, and assigned gene symbols of molecular targets based on annotations in DrugBank^22^ (N=993), articles in PubMed (N=278), Alzforum (N=244), company press releases (N=46), information provided directly in ClinicalTrials.gov submissions (N=19), or other (N=18).

### Human genetic associations

Gene-disease links established by human genetics up through the end of the study period (March 2020) were identified through manual searches of Online Mendelian Inheritance in Man (OMIM)^23^ for Mendelian forms of disease, and Open Targets Genetics (OTG)^24^ for genome-wide association studies (GWAS) of common/complex forms of disease. For GWAS loci mapping to multiple potentially causal genes, the gene with the highest locus-to-gene score^24^ was used; if OTG data were missing or indeterminate, the gene highlighted by the study’s original authors was used.

### Role of the funding source

This work was supported by the National Institutes of Health (R21 TR003040 and R01 NS125255). The funder had no role in study design, analysis, interpretation, or decision to publish.

### Statistics, data, and source code availability

Scripted extraction of ClinicalTrials.gov and DrugBank data used scripts in Python 3.8.9; data analysis and visualization were performed in R 4.2.0. All statistical tests were two-sided and nominal P values less than 0.05 were considered significant. Enrichment analyses used Fisher’s exact test. Tests for temporal trends used linear regression, except for the testing of a temporal trend in disease stage (an ordinal variable), which used ordinal logistic regression (polr from the R MASS package). Loess fits were additionally used for visualization of potentially non-linear temporal trends. Error bars represent 95% confidence intervals of the mean (±1.96 standard errors of the mean). Distributions were compared using Kolmogorov-Smirnov test, which does not assume normality. All data and source code used in this study will be made publicly available at http://github.com/ericminikel/nd_trials and are sufficient to reproduce the figures and statistics herein.

## Results

### Characteristics of neurodegenerative disease trials

Of 4,542 trials returned by our search strategy, 3,241 met inclusion criteria and were manually curated (Figure 1). In order to understand the landscape of neurodegenerative disease trials, we first considered both the simple count of trials (Figure 2A), as well as the total patient-years of enrollment (Figure 2B) as potentially a better proxy for R&D spend, in a univariate breakdown of trials by disease area, sector, intervention type, phase, and control group status. The majority of trials were non-industry-sponsored, lacked a placebo or standard of care (SOC) arm, and for a plurality, phase was other/unspecified. But whereas only a minority of trials were industry-sponsored (N=1,240, 38%), these trials were larger on average, and so accounted for a majority (64%) of all patient-years. Drug interventions accounted for a majority of trials (N=1865; 58%) but an even larger majority of patient-years (76%). In a bivariate cross-tabulation (Figure 2C), drug trials accounted for not only the largest number of trials but were also the most intense (patient-years/trial), especially for Alzheimer’s disease, Phase III, placebo/SOC-controlled, and industry-sponsored. Overall, industry-sponsored drug trials accounted for 61% of all patient-years (Figure 2D). The remainder — non-industry and/or non-drug trials — were highly skewed in terms of size: 34% enrolled ≤20 patients, while the 6 largest trials accounted for 33% of all patient-years, indeed, a single trial of medication adherence reminder devices^25^ that included PD patients comprised 13%. Industry-sponsored drug trials were less variable in size: it took the top 25 to comprise 33% of patient-years, and only 15% off trials enrolled ≤20 patients (Figure 2D). Industry-sponsored drug trials also differed from other trials in being 3.1 times more likely to have a placebo or standard of care (SOC) control arm, 2.2 times as likely to complete, and >100 times as likely to have a specified phase (Figure 2E). Industry-sponsored drug trials grew much more selective over the two decades considered here, with the mean number of inclusion and exclusion criteria per trial rising from ∼9 to ∼17 (P = 3e10, linear regression; Figure 2F), with no corresponding trend for non-industry and/or non-drug trials P = 0.31, linear regression; Figure 2F). The number of inclusion and exclusion criteria rose for Phase I, II, and III trials, but not for other/unspecified phase trials (Figure 2G).

**Figure 2.**
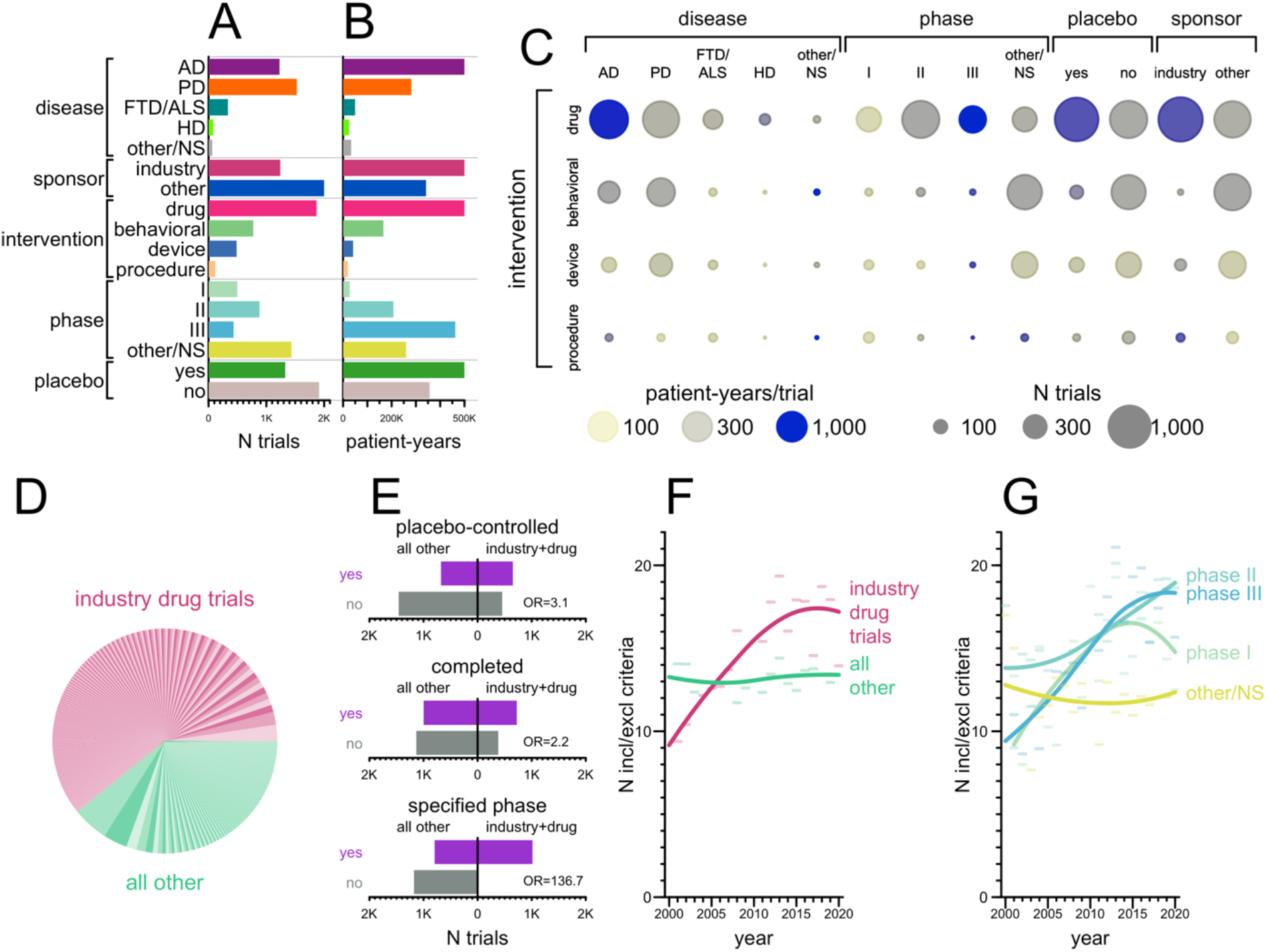
Characteristics of neurodegenerative disease clinical trials. **A)** Univariate count of trials by disease, sponsor, intervention class, phase, or placebo/SOC control status. **B)** Total Univariate total of patient-years (see Methods) by the same variables. **C)** Bivariate cross-tabulation of number of trials and patient-years. Intensity (patient-years per trial) is expressed as a color palette from transparent yellow to opaque blue; number of trials is expressed as the size of each circle. **D)** Pie chart of patient-years of enrollment, with industry drug trials shown in 3 alternating shades of pink and all other trials in 3 alternating shades of green. Each wedge represents one trial, and trials are sorted by number of patient-years. **E)** Barplot representation of contingency tables for whether trials are (purple) or are not (gray) placebo/SOC-controlled (top), completed (middle), or have a specified phase (bottom) depending on whether they are industry-sponsored drug trials (right) or other (left). **F-G)** Mean total number of inclusion and exclusion criteria per trial as a function of (F) industry-sponsored drug trials versus all other, and (G) phase. Loess curves were fit on the raw individual values, but due to the large number of trials, individual values are not shown; instead the average for each year is shown as a semitransparent horizontal bar. Statistical significance was evaluated by linear regression, see Results text.

### Disease stage of participants in trials

Reading the inclusion and exclusion criteria, we manually curated which disease stages, numbered 0-4 (see Methods), were eligible for each trial. 89% of trials required a diagnosis of the neurodegenerative disease in question, corresponding to disease stage 4 (Figure 3A). Another 7.8% permitted patients with mild cognitive impairment (MCI) or an analogous level of other functional impairment, who did not yet meet diagnostic criteria for their diseases (Figure 3A). The 2.7% of trials (N=89) that permitted pre-symptomatic patients, corresponding to stages 0-2 (Figure 3A), differed from other trials in several respects. Trials open to pre-symptomatic individuals were less likely to be industry-sponsored and less likely to test a drug or device; they were much more likely to test a behavioral intervention (Figure 3B). They trended less likely to have a specified phase and had a non-significantly lower completion rate, although the proportion that included placebo/SOC arms was similar. Trials open to pre-symptomatic individuals were significantly longer on average (P = 0.0004, Kolmogorov-Smirnov test; Figure 3C) and trended slightly larger in enrollment, though the difference was not significant (P = 0.09, Kolmogorov-Smirnov test). The proportion of trials enrolling earlier disease stages rose slightly in recent years (P < 1e-10, ordinal logistic regression), although in absolute terms, the proportion of trials enrolling at stages <4 rose only from 8% in the first 4 years of the data to 15% in the final 4 years of data. We also examined whether quantitative measures of impairment changed over time, regardless of nominal disease stage. A majority of trials (57%, N=1,833/3,241) used a disease severity scale as one inclusion or exclusion criterion, most often the Mini-Mental State Examination^20^ (MMSE; 28%, N=911) or Hoehn & Yahr^21^ (23%; N=757). For MMSE, on average, the maximum (least impaired) admissible score rose from 25.6 in 2000 to 27.6 in 2020 (P=2e-5, linear regression), while the minimum (most impaired) admissible score rose from 13.6 to 17.7 (P=1e-5; Figure 3E). Thus, trials using MMSE focused on less impaired patients over time, and because the exclusion of too-advanced patients became stricter more rapidly than the inclusion of less advanced patients, the size of the eligible window shrunk over time. Analogously, for Hoehn & Yahr, where higher scores correspond to more advanced disease, the average minimum (least impaired) admissible score dropped from 1.6 to 1.2 over the twenty years (P=0.002), while the average maximum (most impaired) limit dropped from 3.8 to 3.1 (P=6e-10), again reflecting a shift towards less impaired patients together with a shrinking of the eligible window (Figure 3F).

**Figure 3.**
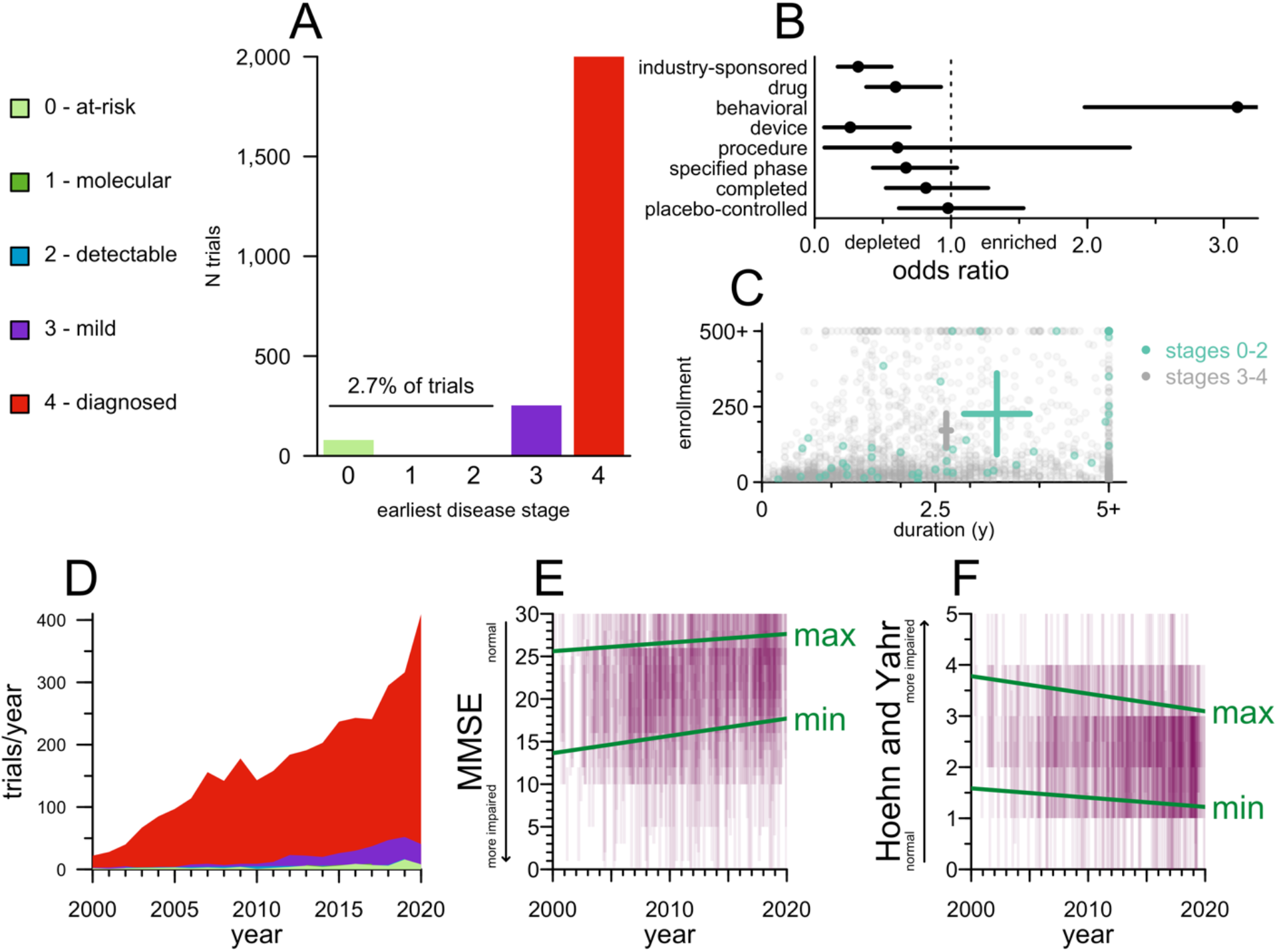
Disease stages eligible for trials. **A)** Barplot of trial count by earliest disease stage (legend at left, see Methods for details) eligible to enroll. **B)** Forest plot of odds ratios (Fisher’s exact test) for properties of preventive (stage 0-2) versus symptomatic (stage 3-4) trials. **C)** Scatterplot of study duration (x axis) and enrollment (y axis) for preventive (stage 0-2, cyan) versus symptomatic (stages 3-4, gray) trials. Crosshairs represent mean and 95% confidence intervals of the mean on both dimensions. **D)** Stacked area plot of the number of trials per year by earliest eligible disease stage. For 2020, only 3 months of data were included, so the raw number of trials was scaled by a factor of 4 to yield trials/year. **E)** Eligible MMSE score ranges by year, N=911. Each trial is displayed as a purple rectangle of 10% transparency stretching from the lowest to highest eligible score on the y axis and staggered by ±0.5 years on the x axis, such that darker shades of purple indicate a greater density of trials recruiting patients in a given score range in a given year. Green lines represent best fits from linear regression models. Lower scores indicate greater impairment. **F)** As in E, but for Hoehn and Yahr, N=757; note that on this scale, higher scores indicate greater impairment.

### Therapeutic hypotheses tested in drug trials

We identified N=750 unique molecular entities (including unique combinations) tested across N=1,865 drug trials. Based on regulatory status and molecular target(s), we classified these trials into 7 categories (Figure 3A). There exist 27 drugs that have full approval and are labeled by FDA specifically for the treatment of the neurodegenerative diseases considered here. Trials in support of these approved drugs, for their approved indications, comprised 18% of patient-years. Only a minority, however, were launched in years prior to FDA approval (Figure 4A). Trials of these same molecular entities occurring after first approval — for example, those seeking to expand the label, meet regulators’ requirements in other countries, understand drug effects on additional endpoints, or test new formulations or delivery routes of the same molecular entity — outnumbered the trials preceding initial approval by a factor of >4 (Figure 4B). Indeed, donepezil, whose approval for Alzheimer’s (1996) pre-dates the time range considered here, was the single most intensely studied drug in this entire dataset (N=56 trials). For each of the drugs approved for these diseases prior to 2017, there were at least as many post-approval as pre-approval trials (Figure 3B).

**Figure 4.**
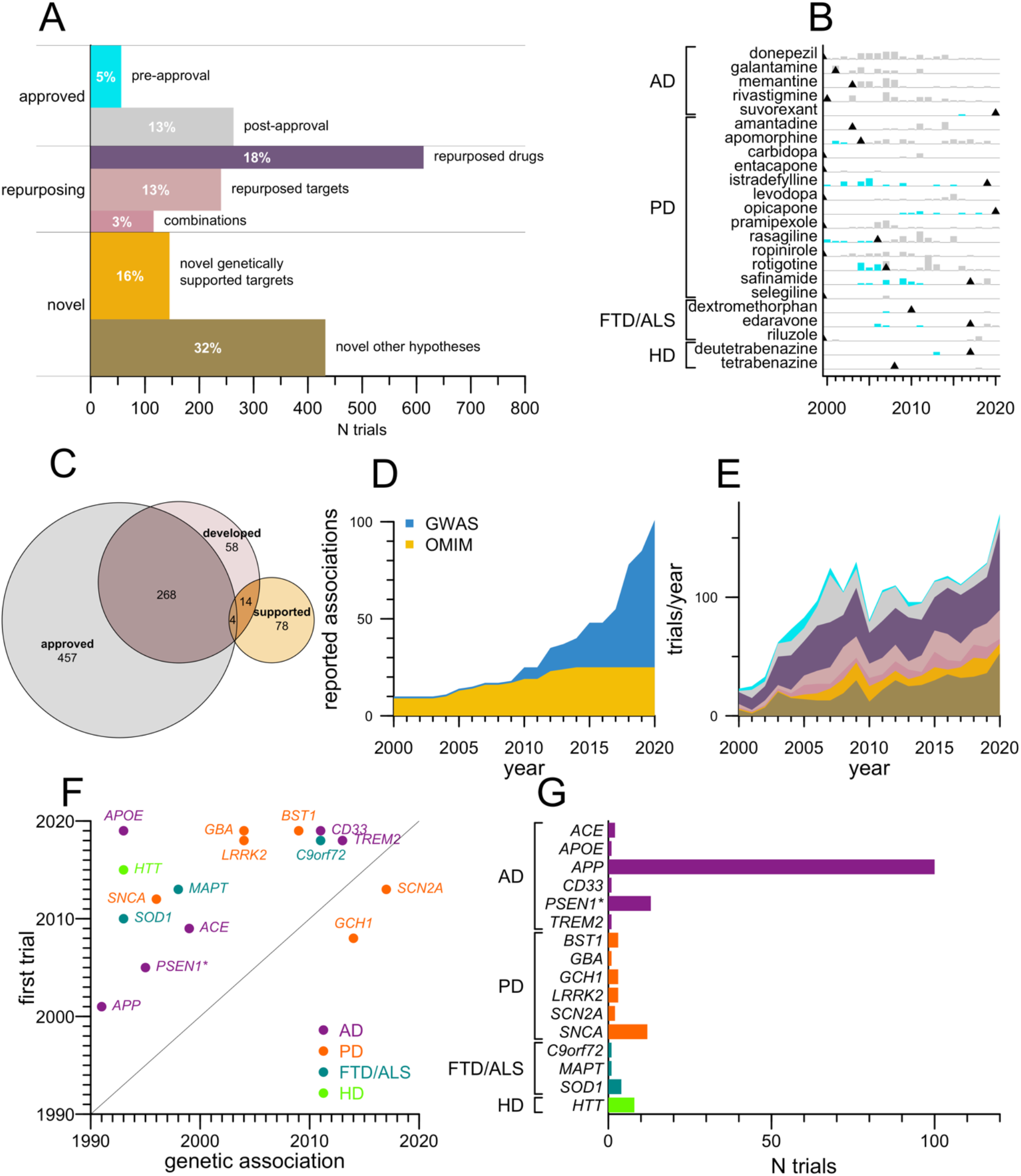
Therapeutic hypotheses tested in drug trials. **A)** Area-scaled barplot of 7 categories of drug interventions tested. The length of each rectangle on the x axis is the number of trials; thickness on the y axis is proportional to the number of patient-years per trial. Thus the total area occupied by each rectangle is proportional to the total patient-years invested in each category, for which percentages of the total are overlaid in white text. **B)** Barplot of number of trials per year for drugs FDA approved for the treatment of the indicated disease before (cyan) and after (gray) initial FDA approval. Trials of these same drugs in other neurodegenerative diseases, for which they are not yet labeled by FDA, are not included in this plot; trials where these drugs served as SOC arms are also excluded. Black triangles indicate years of first FDA approval; approvals prior to 2000 are left-aligned. **C)** Proportional area Euler diagram of genes that encode targets of drugs approved for any indication (gray), targets tested in neurodegenerative disease clinical trials in this dataset (red), or targets supported by human genetic evidence (yellow). **D)** Stacked area plot of cumulative number of genes associated to these 4 diseases by year. Association of ACE to AD is counted in 1999 acknowledging candidate gene studies^26^ which replicated by GWAS in 2018^27^. **E)** Stacked area plot of categories of trials as defined in (A) by year. **F)** Scatterplot of genetically supported target-indication pairs pursued in drug trials, displayed as year of first reported genetic association (x axis) versus year of first trial in the genetically linked neurodegenerative disease (y axis), color-coded by disease. *Gamma secretase is represented here by PSEN1; members PSEN2 and APH1B also have genetic association to AD risk. **H)** Barplot of number of trials for each genetically supported target, in its genetically supported indication, tested clinically.

Another 34% of patient-years were spent on trials of either repurposed drugs, new drugs for established targets (targets with a drug approved for any disease), or combinations of 2 or more therapies where all are either approved drugs or supplements. In all, trials explored 272 different targets of approved drugs (Figure 3C). Of repurposing efforts, 56 trials used a drug approved for one neurodegenerative disease and tested its efficacy in a different neurodegenerative disease (for example, the Alzheimer’s drug memantine was trialed for Parkinson’s, Huntington’s, and FTD/ALS). Ten times as many trials (N=557), however, tested drugs approved for other indications, chiefly in neurology (N=194), metabolic (N=72), and cardiovascular disease (N=46); the most studied repurposed drug was botulinum toxin A (N=17).

Trials comprising 48% of patient-years tested novel therapeutic hypotheses — molecular entities not yet approved and whose molecular targets are either unknown, or are not yet targeted by any other approved drug. Curating associations from both Mendelian forms and genome-wide association studies for these 4 diseases (see Methods), we identified N=101 gene-disease pairs linked by human genetic evidence (Figure 4D). We asked which of these therapeutic hypotheses had been tested clinically. Approximately three-quarters (432/577) of trials of novel therapeutic hypotheses lacked direct human genetic association implicating the target in the disease. The two most intensely studied targets in this group were those with functional evidence for disease relevance (*MAPT* and *BACE1* in AD, N=23 trials each). For the majority of trials in this category (71%, N=307), however, we were unable to identify any known molecular target. The remaining (145/577) trials tested N=16 target-indication pairs backed by human genetics. Their share of all drug trials did not increase over time (P=0.59, linear regression; Figure 4E) despite the increased number of genetic associations reported (Figure 4D). Instead, the only two categories of trial whose share increased significantly were repurposed targets and novel hypotheses without genetic support (P=0.02 and 0.008 respectively). For 14 genetically supported targets, the first clinical trial followed the discovery of the genetic association, with a mean lag time of 13 years, and a minimum lag time of 5 years (*TREM2*; Figure 3F). Trials of genetically supported hypotheses only comprised half as many patient-years as novel hypotheses without genetic support (Figure 3A), and no genetically supported agent was studied as intensely as gingko biloba, to which the two largest trials in the dataset were devoted. Investment in genetically supported target-indication pairs was highly skewed, with 69% of trials and 84% of patient-years devoted to targeting Aβ (*APP*) in Alzheimer’s disease (Figure 3G). Just 3 trials tested genetically supported hypotheses in a preventive paradigm, enrolling individuals at disease stages 0-2: A4^28^, API^29^, and DIAN-TU^30^, all of which tested Aβ antibodies.

## Discussion

Here we used two decades of clinical trial registration data to analyze the characteristics of trials in 4 major neurodegenerative diseases. We were motivated by evidence from other disease areas showing that drug programs whose therapeutic hypotheses are supported by human genetic associations enjoy doubled success rates^15^. We analyzed trial types, disease stages and therapeutic hypotheses being tested to assess to what degree this opportunity has been utilized in major neurodegenerative diseases.

Our findings suggest the risk of a missed opportunity. Most of the genetic studies that have nominated new molecular targets are familial linkage or case-control studies, and thus are best suited to identify the initial triggers of disease. There appears to be an imperfect overlap between the molecular drivers of neurodegenerative disease initiation and the molecular drivers of subsequent progression^31–34^. For some targets nominated by these types of genetic studies, pre-symptomatic populations might represent the best, or in some cases only, opportunity for efficacy. However, trials are overwhelmingly conducted in symptomatic patients. To the extent that trial enrollment has shifted toward less impaired patients over the past two decades, this has come at the expense of screening more patients out, with proliferating inclusion and exclusion criteria and narrower acceptable score ranges. Trials in pre-symptomatic patients remain vanishingly rare, comprising just 2.7% of all trials, and this small fraction is enriched for behavioral interventions and depleted for drug trials and industry sponsorship. Industry-sponsored drug trials, despite representing a minority of all trials in our dataset, accounted for a majority of patient-years of enrollment, and were much more likely than other trials to complete, to have placebo/SOC control arms, and to have a specified trial phase. It is reasonable to conclude that these industry-sponsored drug trials likely represent a large majority of the “shots on goal” for well-powered demonstrations of clinical efficacy.

Trials testing hypotheses rooted in human genetics are a minority and have not become more common despite a proliferation of genetic associations. More common types of trials include those of agents without any known molecular target, post-approval trials of approved symptom-managing drugs, and repurposing trials of drugs approved for other indications. Across the pharmacopeia, of the 729 targets corresponding to drugs approved for any condition, 272 (37%) were tested for neurodegenerative disease, while of 101 hypotheses nominated by human genetics, just 16 were tested. The average time from genetic discovery to first human trial was more than a decade, and the majority of trials and an even larger majority of patient-years focused on Aβ in AD, with limited attention paid to other potential targets.

Our study’s limitations include non-exhaustive capture of trials by ClinicalTrials.gov, the limited amount and types of data available in trial registrations, human error in the curation process, and the inherently retrospective nature of the analysis.

Our analysis suggests that there remain untapped opportunities to explore the disease-modifying potential of genetically validated targets in neurodegenerative disease, but that additional effort to support well-powered, well-controlled trials at earlier disease stages may be needed to realize this potential.

## Data Availability

All data and source code used in this study will be made publicly available at http://github.com/ericminikel/nd_trials and are sufficient to reproduce the figures and statistics herein.

## Notes

### Competing Interest Statement

MAM reports no disclosures. SMV has received speaking fees from Ultragenyx, Illumina, and Biogen, consulting fees from Invitae, and research support in the form of unrestricted charitable contributions from Ionis, Gate, and Charles River. EVM has received consulting fees from Deerfield and research support in the form of unrestricted charitable contributions from Ionis, Gate, and Charles River.

### Summary of Updates

Additional panels in Figure 4, broader review of prior literature in introduction.

